# PET markers of tau and neuroinflammation are co-localized in progressive supranuclear palsy

**DOI:** 10.1101/19010702

**Authors:** Maura Malpetti, Luca Passamonti, Timothy Rittman, P. Simon Jones, Patricia Vázquez Rodríguez, W. Richard Bevan-Jones, Young T. Hong, Tim D. Fryer, Franklin I. Aigbirhio, John T. O’Brien, James B. Rowe

**Affiliations:** Department of Clinical Neurosciences, University of Cambridge, Cambridge, UK; Institute of Molecular Bioimaging and Physiology, National Research Council, Milano, Italy; Department of Psychiatry, University of Cambridge, Cambridge, UK; Cambridge University Hospitals NHS Trust, Cambridge, UK

## Abstract

**Background:** Progressive Supranuclear Palsy (PSP) is associated with tau-protein aggregation and neuroinflammation, but it remains unclear whether these pathogenic processes are related *in vivo*.

**Objectives:** We examined the relationship between tau pathology and microglial activation using [^18^F]AV-1451 (indexing tau burden) and [^11^C]PK11195 (microglial activation) PET in n=17 patients with PSP-Richardson’s syndrome.

**Methods:** Non-displaceable binding potential (BP_ND_) for each ligand was quantified in 83 regions of interest (ROIs). [^18^F]AV-1451 and [^11^C]PK11195 BP_ND_ values were correlated across all ROIs. The anatomical patterns of [^18^F]AV-1451 and [^11^C]PK11195 binding co-localization was determined across sets of regions derived from principal component analyses (PCAs). Finally, PCA-derived brain patterns of tau pathology and neuroinflammation were linked to clinical severity.

**Results:** [^18^F]AV-1451 and [^11^C]PK11195 binding were positively related across all ROIs (r=0.577, p<0.0001). PCAs identified four components for each ligand, reflecting the relative expression of tau pathology or neuroinflammation in distinct groups of brain regions. Positive associations between [^18^F]AV-1451 and [^11^C]PK11195 components were found in sub-cortical (r=0.769, p<0.0001) and cortical components(r=0.836, p<0.0001). PCA-derived components reflecting tau burden (r=0.599, p=0.011) and neuroinflammation (r=0.713, p=0.001) in sub-cortical areas related to disease severity.

**Conclusions:** We show that tau pathology and neuroinflammation co-localize in PSP, and that individual differences in subcortical tau pathology and neuroinflammation are linked to clinical severity. Although longitudinal studies are needed to determine how these molecular pathologies are causally linked, we suggest that the combination of tau- and immune-oriented strategies may be useful for effective disease-modifying treatments in PSP.

## Introduction

Progressive supranuclear palsy (PSP) is a devastating neurodegenerative disorder caused by the neuro-glial aggregation of tau protein, particularly in the basal ganglia, diencephalon, and brainstem (1). The classical clinical phenotype of PSP is the sporadic Richardson’s syndrome, with vertical supranuclear gaze palsy, akinetic-rigidity, falls, and cognitive decline (2). The aggregation of misfolded and hyper-phosphorylated tau protein, first to oligomers and then fibrillary tangles, is central to the PSP pathophysiology (3–6), especially with tau isoforms that have 4 repeats of the microtubule-binding domain (7). Despite the undoubted importance of tau pathology in PSP, neuroinflammation has also been recognized as another ‘core’ aspect in the pathophysiology of PSP (8,9), with a proposed toxic alliance between tau-mediated neurodegeneration and inflammation. Moreover genetic variants linked to inflammatory pathways are overrepresented in PSP *versus* healthy adults (10,11).

We used positron emission tomography (PET) to study the *in vivo* co-localization of tau accumulation and neuroinflammation in PSP. The ligand [^18^F]AV-1451 binds to aggregated tau in Alzheimer’s disease (AD) and, with lower affinity, to non-AD tauopathies like PSP ((12,13);see review (14)). However, [^18^F]AV-1451 does not distinguish tau-from TDP43-pathololgies but this is not problematic in PSP-Richardson’s syndrome because: 1) the clinical-pathological correlation in PSP is very high (15); 2) TDP43 pathology is exceedingly rare in PSP, and 3) [^18^F]AV-1451 displays a specific anatomical pattern of binding that clearly distinguishes PSP from AD (16). Similarly, the ligand [^11^C]PK11195 is widely used as a marker of microglial activation, a proxy of neuroinflammation (17). [^11^C]PK11195 binds to the translocator protein (TSPO) on mitochondrial membranes in activated microglia. In contrast to second generation ligands, it is not significantly influenced by common genetic polymorphisms.

Previous PET studies have confirmed changes in PSP using ligands that target tau (16,18–26) and microglial activation (27,28). However, these studies have not addressed the key issue of whether and how *in vivo* tau pathology and microglial activation are related in PSP. Answering this question would shed light on the pathophysiological mechanisms underlying PSP and may facilitate the development of therapeutic strategies that synergistically target neuroinflammation and tau pathology in PSP.

This study aimed to determine the correlation between microglial activation and tau burden in patients with PSP. We test the hypothesis that neuroinflammation and tau protein aggregation co-localise, and correlate with clinical severity. We assessed the topography of[^11^C]PK11195 and [^18^F]AV-1451 patterns of binding using: 1) a regions of interest (ROI) approach in which non-displaceable binding potential (BP_ND_) values for each ligand were correlated across all ROIs; and 2) a principal component analysis (PCA) which reveals the set of spatially distributed patterns of pathogenic processes, representing the distribution and heterogeneity of pathology in PSP.

## Methods

### Participants

As part of the Neuroimaging of Inflammation in Memory and Other Disorders (NIMROD) study (29), we recruited 17 patients with a clinical diagnosis of probable PSP according to Movement Disorder Society (MDS) 1996 criteria (30). All patients also met the later MDS-PSP 2017 criteria for PSP-Richardson’s syndrome (31). Patients underwent PET scanning with both [^18^F]AV-1451 and [^11^C]PK11195, to respectively assess tau pathology and neuroinflammation. To minimise radiation exposure in healthy people, two groups of control participants were enrolled: n=15 underwent [^18^F]AV-1451 PET and n=16 underwent [^11^C]PK11195 PET. At the first visit, demographic and cognitive measures (i.e., the revised Addenbrooke’s Cognitive Examination - ACE-R) were collected in all participants. Disease severity of patients was measured at the baseline visit and at six monthly intervals, using the PSP rating scale (PSP-RS) (32).

All participants had mental capacity to take part in the study and provided written informed consent. The NIMROD protocol was approved by the National Research Ethic Service’s East of England Cambridge Central Committee and the UK Administration of Radioactive Substances Advisory Committee.

### PET and MRI data acquisition and pre-processing

Full details of the imaging protocol have been published elsewhere (16,28). In brief, patients underwent 3T MRI together with [^18^F]AV-1451 and [^11^C]PK11195 PET, using dynamic imaging for 90 and 75 minutes respectively. MRI used Siemens Magnetom Tim Trio and Verio scanners (Siemens Healthineers, Erlangen, Germany), while PET used a GE Advance and GE Discovery 690 (GE Healthcare, Waukesha, USA). The interval between the patient PET scans had a mean and standard deviation (SD) of 1.18 ± 1.67 months.

For each subject, the aligned dynamic PET image series for each scan was rigidly co-registered to the T1-weighted MRI image. BP_ND_ was calculated in 83 cortical and subcortical ROIs using a modified version of the Hammers atlas (33,34), which includes brainstem parcellation and the cerebellar dentate nucleus. Prior to kinetic modelling, regional PET data were corrected for partial volume effects from cerebrospinal fluid by dividing by the mean regional grey-matter plus white-matter fraction determined from SPM segmentation. For [^11^C]PK11195, supervised cluster analysis was used to determine the reference tissue time-activity curve and BP_ND_ values were calculated in each ROI using a simplified reference tissue model with vascular binding correction (35). For [^18^F]AV-1451, BP_ND_ values were quantified in each ROI using a basis function implementation of the simplified reference tissue model (36), with superior cerebellar cortex grey matter as the reference region. This cerebellar region was selected as reference region given post-mortem evidence showing minimal tau pathology in PSP (see Supplementary material in Passamonti et al. (16)). The same data acquisition and analysis approach was applied for the two control groups.

### Statistical analyses

Age, years of education, ACE-R total and fluency scores were compared between patients and controls with independent-samples t-tests, while gender was compared with the Chi-square test. Linear regression models were applied to the longitudinal PSP-RS scores in individual patients to estimate the PSP-RS score at the time of each PET scan. Image analysis proceeded in four steps. All statistical analyses were performed in SPSS Statistics version 25 (IBM).

First, to test whether microglial activation and tau pathology co-localised across the whole brain, we estimated the Pearson correlation of corresponding [^11^C]PK11195 and [^18^F]AV-1451 BP_ND_ group-average values across all 83 ROIs (Figure 1).

**Figure 1.**
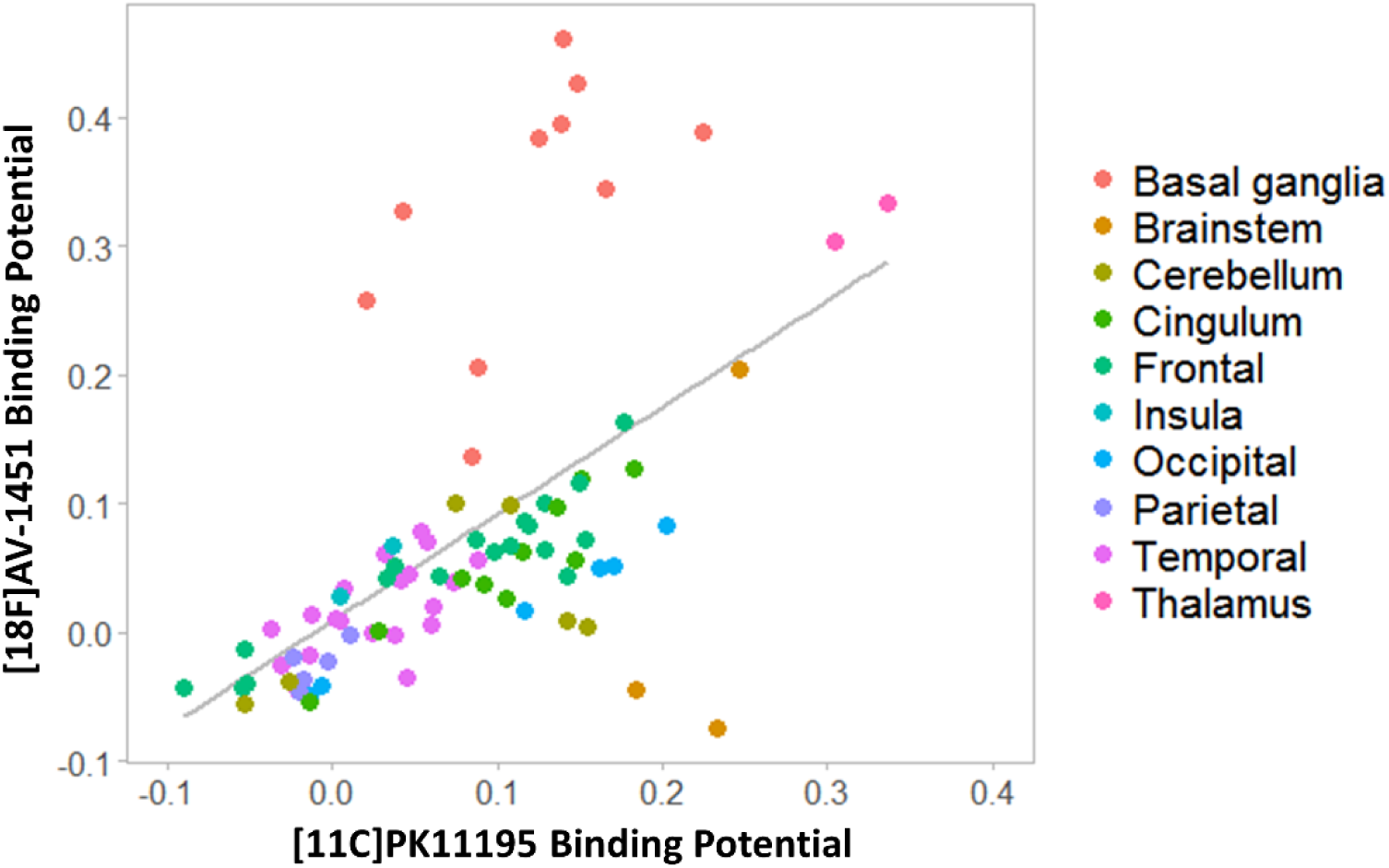
Whole brain correlation between regional mean non-displaceable binding potential (BP_ND_) of [^11^C]PK11195 and [^18^F]AV-1451 in the PSP group. Each point represents the average value across all patients for a specific brain region, while colours indicate brain macro-areas.

Second, the number of ROIs was reduced from 83 to 46, averaging left and right regional BP_ND_ values, as in previous studies (16,28). This step reduces the degrees of freedom, increasing power, and is justified in PSP in which the motor syndrome is essentially symmetric. The differences between PSP and control groups in the 46 ROIs were tested for each ligand with independent t-tests with false discovery rate (FDR) correction for multiple comparisons.

Third, in PSP patients, BP_ND_ values in the 46 bilateral ROIs were included in separate PCAs for [^11^C]PK11195 and [^18^F]AV-1451. This reduces the data dimensionality further, identifying a small set of components that best explain the data variance which. The resulting component reveal anatomical patterns covary in terms of neuroinflammation or tau pathology. Varimax rotation was applied in the PCA to increase orthogonality across the different components (i.e., anatomical patterns of neuroinflammation and tau pathology). The components with eigenvalues > 1 were retained, explaining >80% of the cumulative variance.

Finally, to test for co-localization of microglial activation and tau pathology in specific neuroanatomical patterns of ligand binding, we performed Pearson correlations between individual scores of each ligand-specific component extracted. The analyses adjusted for age differences and variability in the time interval between PET scans, included as covariates of no interest. For each ligand, we tested for correlations between regionally specific PCA clusters (i.e., anatomical patterns of neuroinflammation and tau pathology) and disease severity (the estimated PSP-RS score at the time of each scan). Bonferroni’s method was used to correct for multiple comparisons.

## Results

Demographics, clinical and cognitive variables are summarized in Table 1. There were no group differences in age (t(46)=0.14, p=0.892) or sex (X^2^(1)=0.230, p=0.632). Differences between PSP and control groups were found in the ACE-R total scores (t(46)=4.53, p<0.001), fluency ACE-R sub-scores (t(46)=6.11, p<0.001), and years of education (t(46)=3.64, p=0.001). Mean and SD of estimated PSP-RS scores at the time of PET are also included in Table 1.

**Table 1.** Demographic and clinical characteristics for patients and controls groups (*=significant t-test between groups). Estimated PSP rating scale (PSP-RS) score refers to the adjusted value to the time midway between the two PET scans.

|  | **PSP patients** | **Controls** | **Difference** |
| --- | --- | --- | --- |
| **N** | 17 | 31 |  |
| **Sex**<br>(F/M) | 7/10 | 15/16 | $\chi^2=0.23$, $p=0.632$ |
| **Age**<br>(mean ± SD) | 68.3±5.7 | 68.6±7.1 | $t=0.14$, $p=0.892$ |
| **Education**<br>(mean ± SD) | 12.2±1.9 | 14.8±2.6 | $t=3.64$, $p=0.001^*$ |
| **ACE-R**<br>(mean ± SD) | 82.7±10.5 | 94.6±4.0 | $t=4.53$, $p<0.001^*$ |
| **Fluency**<br>(mean ± SD) | 7.0±3.2 | 12.1±1.7 | $t=6.11$, $p<0.001^*$ |
| **Estimated PSP-RS**<br>(mean ± SD) | 41.6±12.0 | - | |

### Regional [^11^C]PK11195 and [^18^F]AV-1451 BP_ND_ in PSP

Regional group mean [^11^C]PK11195 BP_ND_ correlated with the corresponding [^18^F]AV-1451 BP_ND_ across the whole brain (r=0.577, p<0.0001) (Figure 1). At the group level, patients with PSP had high [^11^C]PK11195 BP_ND_ in the brainstem, cerebellum, thalamus, and occipital and cingulate cortex, with pons and medulla having the highest values (pons: 0.19 ± 0.08; medulla: 0.23 ± 0.13). High [^18^F]AV-1451 BP_ND_ was found in the basal ganglia, midbrain and thalamus (Figure 1), with the basal ganglia having the highest value (mean=0.33; SD =0.10).

[^11^C]PK11195 binding values were higher in PSP patients than in controls in the putamen (t(31)=4.01, p<0.001), and pallidum (t(31)=3.72, p=0.001). Likewise, [^18^F]AV-1451 binding was significantly increased in PSP, than controls, in the putamen (t(30)=3.66, p=0.001), pallidum (t(30)=5.69, p<0.001), thalamus (t(30)=3.74, p=0.001), midbrain (t(30)=3.84, p=0.001), and dentate nucleus (t(30)=3.87, p=0.001), confirming our previous findings (16,28). All comparisons reported survived at the FDR correction (p=0.001 corresponds to p=0.0215 FDR for [^11^C]PK11195 comparisons and p=0.0086 for [^18^F]AV-1451 comparisons). We do not further discuss this group comparison as the principal aim of this study was to study the co-localization of [^11^C]PK11195 and [^18^F]AV-1451 binding in PSP.

### Principal component analysis of [^11^C]PK11195 and [^18^F]AV-1451 BP_ND_ in PSP

For [^11^C]PK11195 BP_ND_, four components were identified which collectively explained 81.4% of the data variance (Figure 2, left panel). Component 1 reflected [^11^C]PK11195 binding in posterior cortical regions, the orbitofrontal cortex and cerebellar grey-matter (62.9% of the total variance). Component 2 grouped together medial and superior regions of the temporal lobe including the amygdala, hippocampus and para-hippocampal gyrus, as well as other cortical areas such as the insula and temporo-parietal junction (9.2% variance). Component 3 was weighted to brainstem regions (i.e., midbrain and pons), the dentate nucleus, and the cerebellar white-matter (5.1% variance). Component 4 comprised superior and medial frontal regions (4.3% variance).

**Figure 2.**
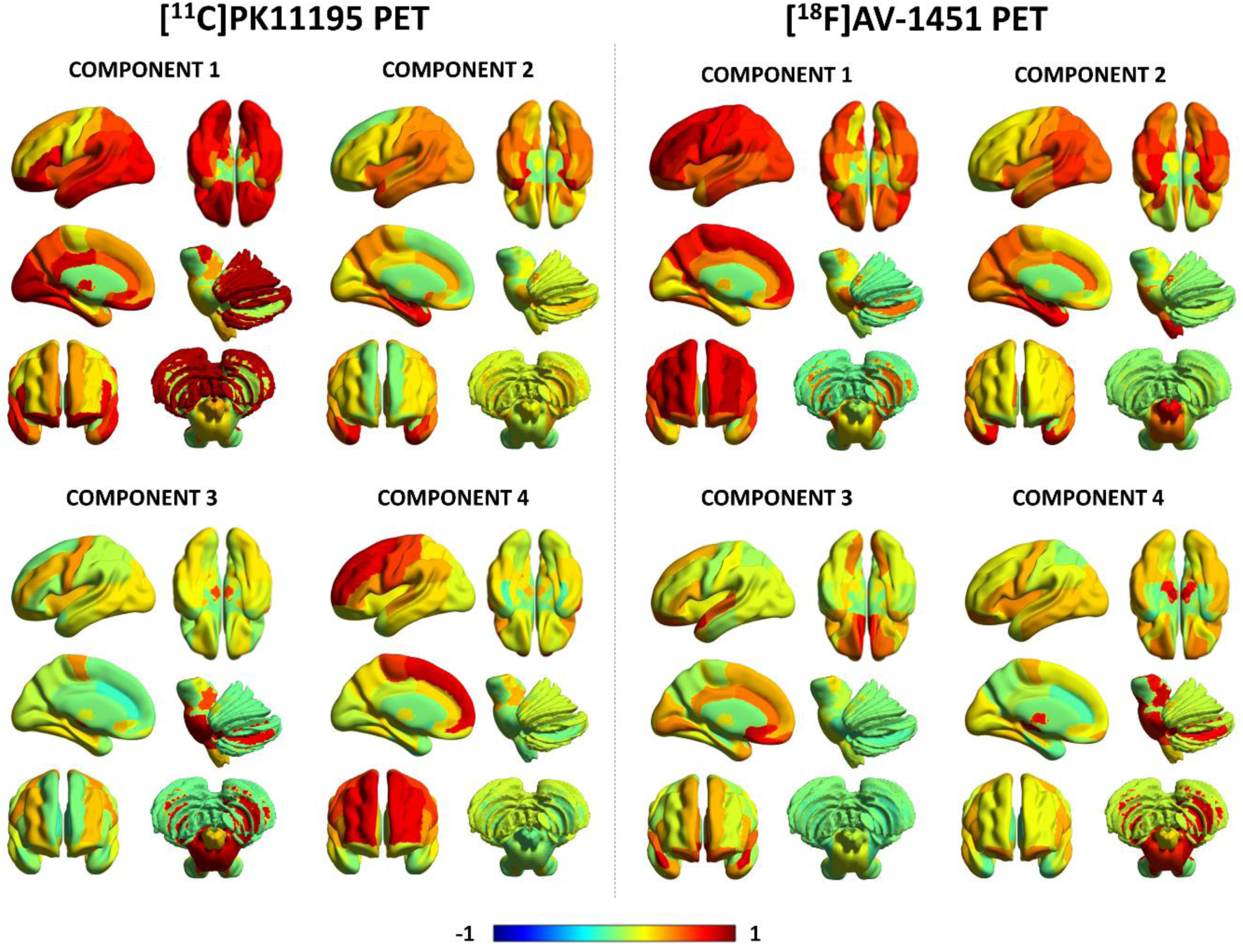
First four principal components for [^11^C]PK11195 non-displaceable binding potential (BP_ND_) and [^18^F]AV-1451 BP_ND_ in the PSP group. The colours represent the rotated weights of all brain regions for each component.

Likewise, four components were found for [^18^F]AV-1451 BP_ND_ which explained together 81.8% of the data variance (Figure 2, right panel). Component 1 reflected global [^18^F]AV-1451 cortical binding, especially in frontal cortical regions (61.3% of the total variance). Component 2 reflected [^18^F]AV-1451 BP_ND_ binding in the insula and medial temporal lobe regions (e.g. amygdala, hippocampus) (8.6% variance). Component 3 loaded onto the anterior superior temporal gyrus and frontal subgenual cortex (7.0% variance). Component 4 was weighted towards subcortical areas including the midbrain, pons, substantia nigra, thalamus, dentate nucleus and cerebellar white matter (5.0% variance).

### Correlation between [^11^C]PK11195 and [^18^F]AV-1451 principal components in PSP

After adjusting for Bonferroni correction (p=0.05/16 correlations between [^18^F]AV-1451 and [^11^C]PK11195 components), Pearson correlations between individual loading scores for the four [^11^C]PK11195 components and the four [^18^F]AV-1451 components showed two significant results (Figure 3): 1) [^11^C]PK11195 component #2 positively correlated with [^18^F]AV-1451 component #2 (r=0.836, p<0.0001), and 2) [^11^C]PK11195 component #3 positively correlated with [^18^F]AV-1451 component #4 (r=0.769, p<0.0001). Importantly, these correlations remained significant after correcting for age differences and variability in the time interval between PET scans ([^11^C]PK11195 #2 - [^18^F]AV-1451 #2: r=0.787, p=0.001; [^11^C]PK11195 #3 - [^18^F]AV-1451 #4: r=0.791, p<0.0001). [^11^C]PK11195 component #1 also weakly negatively correlated with [^18^F]AV-1451 component #1 (r=-0.564, p=0.018, uncorrected).

**Figure 3.**
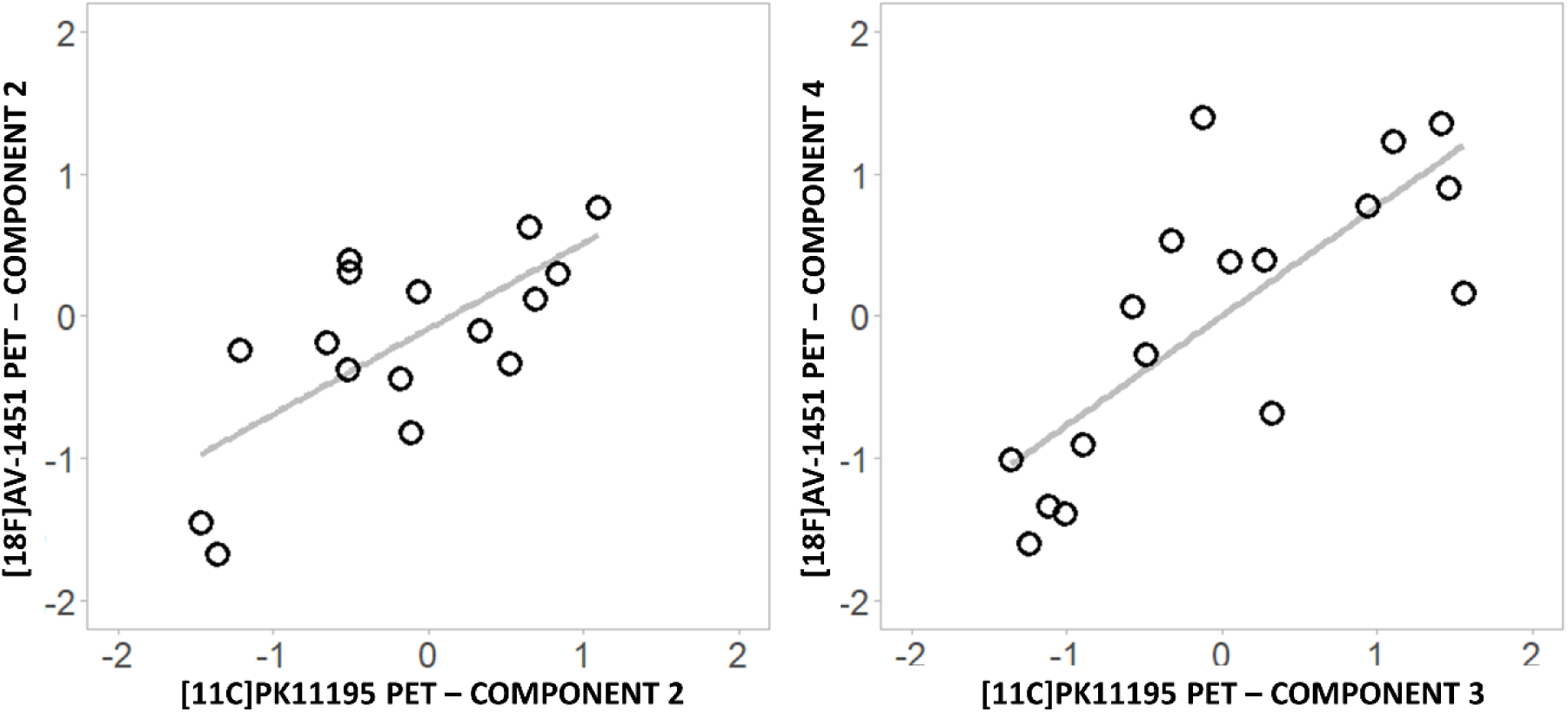
Significant correlations between [^11^C]PK11195 and [^18^F]AV-1451 individual principal component scores.

The individual PCA-derived scores for ligand-specific subcortical components separately correlated with disease severity (PSP-RS) (Figure 4). Both [^11^C]PK11195 component #3 (r=0.713, p=0.001) and [^18^F]AV-1451 component #4 (r=0.599, p=0.011) were positively associated with disease severity as expressed by the estimated PSP-RS values. We then applied a linear model to test whether the interaction between [^11^C]PK11195 component #3 and [^18^F]AV-1451 component #4 predicts PSP-RS (adjusted to the time midway between the PET scans). The model with the interaction between the two components was significant (F(13)=4.571, adjusted R^2^=0.51, p=0.02, AIC=79.11) but the interaction term itself was not (p=0.565). Full backward elimination revealed the best minimal model (F(15)= 14.13, p=0.0019, adjusted R^2^=0.49, AIC=76.07) with only one predictor, [^11^C]PK11195 component #3 (p=0.0019).

**Figure 4.**
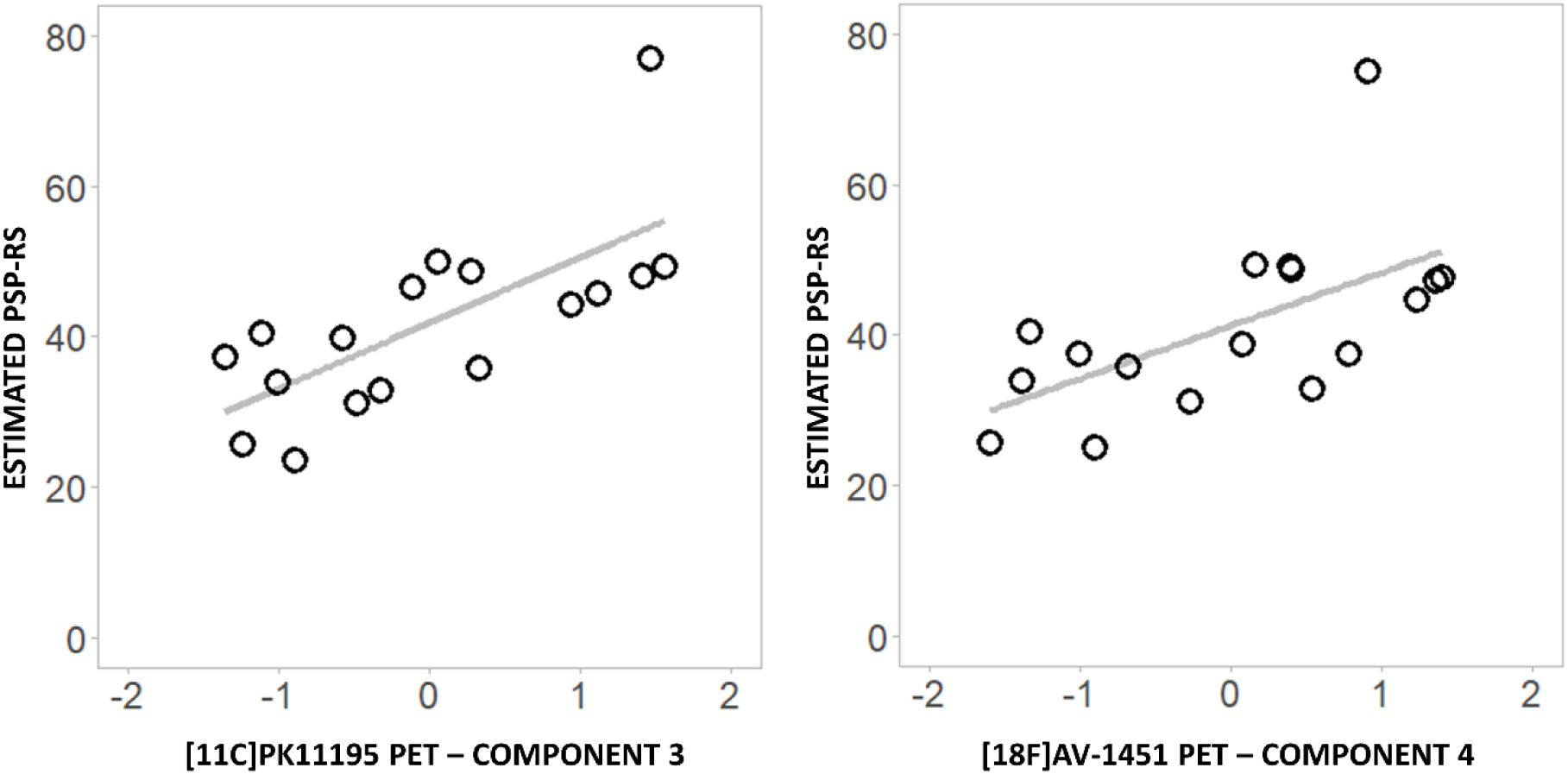
Significant correlations between [^11^C]PK11195 (left) and [^18^F]AV-1451 (right) individual principal component scores and estimated PSP-RS at the time of the two PET scans.

## Discussion

This study suggests that tau protein aggregation and microglial activation are anatomically co-localized in PSP, and relate to disease severity. The relationship is observed across widespread brain regions although it is most evident in some cortical (i.e. insula and temporo-parietal junction) and subcortical regions (i.e. brainstem and cerebellum). The *in vivo* measures of neuroinflammation and tau burden in the brainstem and cerebellum were also individually associated with disease severity.

Before considering the insights of our findings into the pathogenesis of PSP, we discuss the caveats related to the two ligands. Although [^18^F]AV-1451 shows strong *in vivo* and post-mortem binding to tau pathology in AD, it displays a variable affinity in healthy aging and non-AD tauopathies (37–39). This tracer binds to non-tau proteinopathies (i.e. is positive in TDP43 pathology related to C9orf72 mutations and semantic dementia (12)), and nonspecific sites such as neuromelanin (38), monoamine oxidase (MAO; (40)) and choroid plexus (37). However, previous post-mortem data (16) have shown that off-target binding to neuromelanin cannot be the cause for [^18^F]AV-1451 binding in the striatum or the cortex, as these regions do not accumulate neuromelanin (41). In the basal ganglia MAO-A is significantly expressed, and this has been proposed as an alternative ‘off-target’ binding site of [^18^F]AV-1451 (40). MAO-B has also been found to be expressed in reactive astrocytes and activated microglia (40,42), which raises the critical issue of whether [^18^F]AV-1451 binding relates not only to tau pathology but also neuroinflammation. However, this hypothesis is rejected by data in a carrier of a MAPT genetic mutation in which high [^11^C]PK11195 binding in frontotemporal regions was found despite a lack of [^18^F]AV-1451 binding (43). Finally, although [^18^F]AV-1451 binding in the choroid plexus has been described as molecular ‘off-target’ binding in healthy aging, histological analyses have challenged this hypothesis, reporting tangle-like structures in epithelial cells of this region (44).

Nevertheless, we acknowledge that the affinity of [^18^F]AV-1451 to the 4-repeat tau in non-AD tauopathies as PSP is lower than its affinity to 3/4-repeat AD-related tau pathology (38,39). In PSP patients, increased [^18^F]AV-1451 binding has been shown sub-cortical rather than cortical regions, consistently with previous neuropathological evidence (16,18,19,23,24,26). This evidence supports the use of [^18^F]AV-1451 PET to quantify and localise tau pathology in tauopathies with clear and known pathologic substrates, such as PSP. Our previous study (16) also showed that it is possible to discriminate, with machine-learning approaches and multivariate pattern analyses, the neuroanatomical pattern of [^18^F]AV-1451 binding in PSP from the one seen in AD. This corroborates the use of [^18^F]AV-1451 PET as a marker of disease-specific pathological changes.

[^11^C]PK11195 has been criticised for its relatively low signal to noise ratio and low brain penetration which can affect its sensitivity to activated microglia. However, this would only reduce the effect sizes and increase type II error, rather than leading to false positive findings. Several second-generation PET radioligands for TSPO are characterised by higher signal-noise ratio than [^11^C]PK11195 but their binding is affected by single nucleotide polymorphisms (rs6971) that cause heterogeneity in PET data and requires genetic screening (45). In contrast, [^11^C]PK11195 binding is not affected by this polymorphism (45), and it has well established methods of kinetic analysis (35). Hence, [^11^C]PK11195 PET remains the most used method to study microglia activation in neurodegenerative diseases (17), and it has been successfully applied in PSP (27,28).

With these caveats in mind, we now discuss our principal results. To study the *in vivo* co-localization between microglial activation and tau pathology in PSP, we applied correlation analyses between the binding of the two ligands 1) across all brain regions, and 2) between principle components of set of bilateral brain regions, extracted to reduce the complexity of the imaging data. With the first approach, we found a positive correlation between [^18^F]AV-1451 and [^11^C]PK11195 binding, across the whole brain (Figure 1). This indicates a close association between microglial activation and tau pathology in PSP that extensively involves both subcortical and cortical regions. This finding also aligns with *in vivo* correlation between neuroinflammation and tau aggregation in AD and frontotemporal dementia (46,47). Collectively, these multi-tracer PET studies support previous *in vitro* evidence of the association between microglial activation and tau aggregation in different tauopathies (see review (48)). The spatial distribution of the *in vivo* association between microglial activation and tau pathology in PSP also mirrors previous findings about neurodegeneration and tau pathology affecting not only subcortical but also cortical regions in PSP Richardson’s syndrome (6,49).

When assessing the ligand-specific components from principal component analysis, we found a positive correlation between [^18^F]AV-1451 and [^11^C]PK11195 binding in brainstem and cerebellar regions, loaded into anatomically overlapping components of both ligands (component #3 for [^11^C]PK11195 and component #4 for [^18^F]AV-1451) (Figure 3, right panel). This association occurs in motor-related regions that are involved in the neuropathology and symptomatology in PSP (i.e. functional deficits, postural instability, and supranuclear gaze palsy) associated with PSP Richardson’s syndrome (50). Furthermore, both tau pathology and microglial activation in the brainstem-cerebellar component correlated with disease severity, reflected by estimated PSP-RS scores at the time of the PET scans (Figure 4). This finding adds relevant information to the literature as only a few previous studies have explored how [^18^F]AV-1451 and [^11^C]PK11195 binding individually correlate with disease severity in PSP. In three studies, PSP-RS scores did not correlate with [^18^F]AV-1451 uptake in any brain region (16,18,26), while two other studies found a weak correlation in the globus pallidus (23), midbrain, cerebellum and frontal cortex (24). For [^11^C]PK11195, only one study explored the association between ligand binding and clinical severity, finding a significant correlation in the pallidum, midbrain and pons (28). In our sample, although the two components individually correlate with clinical severity, they do not interact in their association with PSP-RS. This suggests an additive and partially independent effect of the two pathological processes on the clinical progression rather than a synergistic effect, although a larger sample size and longitudinal design will be needed to further explore this relationship.

[^18^F]AV-1451 and [^11^C]PK11195 binding were also correlated in a cortical component (#2 for both ligands - figure 3, left panel), which for both ligands was weighted towards regions of the medial temporal lobe, insula and temporo-parietal junction. Previous studies have implicated the medial temporal lobe and limbic structures in basic emotional recognition, which has in turn been found to be impaired in PSP, alongside theory of mind and social cognition (51,52). The recognition of happiness was reported to be preserved in patients with PSP, while the recognition of negative emotions (i.e. anger, disgust, surprise, fear and sadness) was affected in these patients (51). Basal ganglia, insula and amygdala have been reported to be implicated in the recognition of predominately negative emotions, and to be pathologically affected in PSP (49,52,53). The association between microglial activation and tau pathology that we found in limbic regions may complement the biological explanation of emotion-related and social deficits in PSP, however, longitudinal studies are needed to clarify the timing of these interacting effects on the pathological and clinical disease progression.

In addition, our finding of tau and neuroinflammation co-localization in the cortex of patients with PSP Richardson’s syndrome is in keeping with previous post-mortem evidence showing tau pathology and atrophy not only in subcortical and limbic regions, but also in the parietal lobe (49,54). Specifically, the supramarginal gyrus has been described as the most affected brain region in two independent pathological cohorts of patients with PSP Richardson’s syndrome (49,54). The absence of *in vivo* evidence about supramarginal atrophy in the literature may enhance the importance of the association between neuroinflammation and tau accumulation in this region as an early biomarker of a later-stage neuronal loss.

Our study has some limitations. First, we acknowledge the limited power of the analyses related to the relatively small size of our sample. Although our cohort is larger than many previous multi-tracer PET studies on rare neurodegenerative diseases like PSP, our findings will benefit from larger and independent replication samples. Second, the recruitment was based on clinical diagnosis, which was confirmed at each follow-up visit; however, post-mortem pathological confirmation was available for only 7 patients. Third, our results are based on a cross-sectional design, which cannot be used to infer causal relationship between tau and microglial activation. A longitudinal assessment of tau burden, microglial activation and clinical progression in the same individuals alongside mediation analyses are the necessary next steps to clarify the interplay between the two pathological processes and their effect on disease severity across time. Finally, to minimise radiation exposure in healthy individuals, controls were divided in two groups, one of which underwent [^18^F]AV-1451 PET and the other one [^11^C]PK11195 PET. For this reason, a direct comparison of the correlation between the two ligands in patients compared to controls was not possible.

To conclude, our results confirm the relevance of neuroinflammation to PSP-Richardson’s syndrome and a close association with tau pathology in the core regions for this disease. Our findings indicate a topographical overlapping between neuroinflammation and tau pathology in those regions previously described by post-mortem studies as mainly involved in PSP pathophysiology. Although we cannot infer the causal direction in the relationship between pathological mechanisms, we speculate that microglial activation may be activated by an initial tau misfolding and contribute to tau pathology and propagation. The latter, in turn, may lead to further neuroinflammation, as previously suggested in AD (see review (48)). A growing literature from pre-clinical research suggests that microglial activation may precede the formation of NFT (55) and then drive the spreading of pathological tau (56). Our findings suggest that the co-localization of neuroinflammation and tau pathology is an important pathogenetic mechanism in PSP, and both processes may be involved in defining the PSP clinical severity. A better understanding of the interaction between the pathological substrates in PSP and its effects on disease progression may crucially contribute to improving patients’ stratification and clinical trials. Specifically, our results encourage the application of [^18^F]AV-1451 and [^11^C]PK11195 PET as markers of co-localised pathological mechanisms in PSP to develop new targeting therapies and empower clinical trials.

## Data Availability

Anonymized data may be shared by request to the senior author from a qualified investigator for non-commercial use (data sharing is subject to participants’ prior consent to data sharing).

## Acknowledgments

We thank our participant volunteers for their participation in this study, and the radiographers/technologists at the Wolfson Brain Imaging Centre and Addenbrooke’s PET/CT Unit, and the research nurses of the Cambridge Centre for Parkinson-plus for their invaluable support in data acquisition. We thank the East Anglia Dementias and Neurodegenerative Diseases Research Network (DeNDRoN) for help with subject recruitment, and Drs Istvan Boros, Joong-Hyun Chun, and other WBIC RPU staff for the manufacture of the radioligands. We thank Avid (Lilly) for supplying the precursor for the production of [^18^F]AV-1451 used in this study. This study was funded by the National Institute for Health Research Cambridge Biomedical Research Centre and Biomedical Research Unit in Dementia (NIHR, RG64473); the PSP Association; the Wellcome Trust (JBR 103838); the Cambridge Trust & Sidney Sussex College Scholarship (MM); the Medical Research Council (LP: MR/P01271X/1); and the Cambridge Centre for Parkinson-Plus (SPJ, TR).

JBR serves as editor to Brain, and is a non-remunerated trustee of the Guarantors of Brain and the PSP Association (UK). He provides consultancy to Asceneuron, Biogen, UCB and has research grants from AZ-Medimmune, Janssen, Lilly as industry partners in the Dementias Platform UK. JOB has provided consultancy to TauRx, Axon, Roche, GE Healthcare, Lilly and has research awards from Alliance Medical and Merck.

